# Addressing the Unique Needs of Emerging Male Adults in HIV Prevention in Rural Kenya. A Qualitative Study

**DOI:** 10.1101/2025.03.05.25323453

**Authors:** Augustine Kiplagat Bingat, Peninnah M. Kako, Dinah J. Chelagat, Seok Hyun Gwon, Jake Luo

**Affiliations:** Associate Clinical Professor at Bon Secours Memorial College of Nursing, Richmond, Virginia, USA; Professor at College of Nursing, University of Wisconsin-Milwaukee, WI, USA; Senior Lecturer and Dean, Moi University, School of Nursing; Associate Professor at College of Nursing, University of Wisconsin-Milwaukee, WI, USA; Associate Professor and Health Care Informatics Graduate Program Director at College of Health Sciences, University of Wisconsin, Milwaukee, WI, USA

**Keywords:** **HIV**: Emerging male adults, Sub-Saharan Africa, rural settings, Adolescents and Young Adults (AYAs), HIV prevention

## Abstract

Emerging male adults continue to be disproportionately affected by HIV compared to other age groups in Sub-Saharan Africa (SSA). Recent demographic data indicate that AIDS related illness is currently the leading cause of death among young people in SSA. Emerging adulthood is critical when it comes to sexuality because it is marked by the formation of identity and the establishment of more mature and intimate relationships which might increase vulnerability to sexually transmitted infections including HIV. Understanding HIV prevention and testing needs in emerging male adults in rural settings is essential to developing prevention efforts.

This descriptive qualitative study was conducted to understand the HIV prevention and testing needs for emerging male adults in rural Kenyan setting. 30 in-depth interviews and 3 FGDs were conducted with emerging adults in rural Ainabkoi sub-county in Uasin Gishu county in Kenya. Findings pointed out that emerging adults in rural settings experience unique challenges in HIV testing and prevention influenced by socio-cultural, economic, political, and legal factors elevating their risk to HIV infection compared to other age groups.

The study underscore that HIV is still the greatest threat among emerging adults in SSA and will require innovative approach to develop and implement youth and young adults’, especially males, sensitive interventions across multiple sectors that influence HIV prevention knowledge, service use, and treatment options for youths.

**Author Summary:** In our study, we explored the challenges of preventing HIV among young adult men in rural Kenya, a region where traditional health services often struggle to reach effectively. I, along with my colleagues, conducted interviews and discussions to understand what these young men need and how they perceive existing HIV prevention efforts.

We found that factors like limited healthcare access, societal norms, and economic conditions significantly influence their risk and ability to receive proper HIV education and services. The young men expressed a strong desire for more accessible and relevant HIV prevention strategies that respect their unique cultural and social settings.

Our research underlines the importance of creating tailored HIV prevention programs that go beyond conventional methods. These programs should engage with the community’s specific needs and leverage local resources to enhance effectiveness. By focusing on these areas, our work contributes to a broader understanding of how to tackle HIV prevention in similar rural settings globally, suggesting that interventions need to be as diverse as the populations they aim to serve. This approach could inform future strategies not only for HIV but also for other health issues faced by isolated communities.

## 1.0. Introduction

Globally, HIV and AIDS is the leading cause of death among young people and the leading cause death among young people (aged 15-24) in Africa (1,2). Despite the progress made in the past 10 years, with a 46% decline in new HIV infections among young people, the world, death among young people (aged 10-24) in SSA is still behind on achieving the targets set for young people (1). This is a serious public health issue which need urgent cost-effective interventions by local and international players to save the emerging adults from the HIV pandemic.

The Kenya Ministry of Health data shows that, nationally, approximately 29% of all new HIV infections are among adolescents and young people who still bear the greatest impact of the HIV epidemic due to limited access to information, services, stigma and discrimination. As per 2020 statistics, the HIV prevalence in Uasin Gishu country is currently at 5.6% above the national average of 4.3% and prevalence has been on the upward trend in since 2013 which was at 4.3% (3). In Uasin Gishu, the young people aged 15 to 24 make up more than half of the new HIV infections, with the biggest driver of HIV infections being the drug abuse and transactional sex, especially in informal settlements (3,4).

There is limited research on HIV prevention and testing needs of adolescents and emerging male adults and the few available target the young people living in urban towns and cities (2,5,6). However, existing studies primarily focus on urban settings, leaving a significant knowledge gap regarding HIV prevention barriers among rural emerging male adults (18–24).The HIV infection in emerging adult male population is underreported because they have lower rate of HIV testing compared to female counterparts due to structural barriers such as stigma, lack of healthcare access, and limited digital literacy (5). As a result, targeted HIV testing and prevention interventions must be designed specifically for this population. Encouraging men to get tested and treated is a major challenge that is insufficiently addressed in existing research (2), particularly in rural settings where healthcare access and digital interventions remain limited.

Several studies have reported that men in sub-Saharan Africa are less likely to be self-aware of their HIV status compared to their female. For example, a survey conducted in 2016-17 in Tanzania showed that only 45% of men living with HIV (MLWH) were aware of their positive HIV status (7). Programmatic efforts should account for this disparity and recognize that it may be necessary to seek out men for HIV prevention, testing, care in order to eradicate HIV and achieve HIV free generation (5,8).

### 1.2. Theoretical Framework

The theoretical and conceptual framework for the study was first guidance by Modified Social Ecological Model (MSEM) to help visualize multilevel domains of HIV infection risks and guide the development of prevention strategies in emerging adults (9). Secondly the concepts from theory of development of emerging adults from the late teens through the twenties proposed by Arnett were utilized (10).

## 2.0. Materials and Methods

The descriptive qualitative design guided the study.

### 2.1. Study Design

Descriptive qualitative research design was used for this study to understand HIV prevention and testing needs for emerging male adults in rural setting in Kenya.

### 2.2. The Sample and Setting

The study sample consist of 60 male participants aged between 18-24 years. A total of 30 participants participated in the individual interviews and another 30 were engaged in the Focus Group Discussions (FGDs). The recruitment and interviewing of participants took place over 6 weeks in spanning between July-August 2021. Purposive sampling enhanced with snowball sampling was used to recruit participants. These methods were necessary due to the difficulty in accessing this demographic through traditional means. We minimized selection bias by recruiting participants through multiple community networks and healthcare outreach programs, ensuring broad representation. These sampling methods assured that the participants will provide insightful information that enhances the understanding of their experience about HIV prevention risk factors and needs unique to this age group.

The study was conducted in largely rural Ainabkoi Sub-County of Uasin Gishu County in Kenya, East Africa. Uasin Gishu County is one of the 47 Counties of Kenya and is in located in the former Rift Valley Province. The town of Eldoret is the county’s largest population center as well as its administrative and commercial center. The county has an estimated population of 894,179 with rural population contributing about 69% of the entire population (11).

### 2.3. Recruitment

The recruitment and interviewing of participants took place over 6 weeks spanning July to August 2021. In the process of recruiting the participants for individual and FGDs, the initial contact was made through local community leaders at Ainabkoi sub-country administrative offices. The focal persons then recommended participants to be contacted for interviews, based on experience of working with the young people in their area of jurisdiction. The purpose of the study was explained to the community leaders and seek their cooperation and help in identifying potential participants.

### 2.4. Data Collection

Data was collected from individual, focus groups and key informants.

#### 2.5.1. Key Informants Interview

The Ainabkoi Sub-country HIV and AIDS coordinator was interviewed about HIV and AIDS prevalence, policies and prevention programs targeting the young people in the county and sub-counties. This method was utilized to get data about higher-level factors namely policy and state of HIV epidemic in Uasin Gishu County and Kenya in general.

#### 2.5.2. Individual Interviews

Individual interviews were conducted face-to-face by the investigator in a private space/room in different localities in Ainabkoi Sub-County. The interviewer opened the discussion with general questions and went ahead to discuss the participant’s HIV risks factors and needs in HIV testing and prevention unique to this age group. The duration of each interview was about 60 minutes. Interviews was audio recorded and the participants were not identified by their numbers but serial numbers on the tape to maintain confidentiality.

#### 2.5.3. Focus Group Discussions (FGDs)

There were three follow-up FGDs and participants were composed of different individuals from the in-depth interviews. Conducting the three follow-up FGDs was for triangulating and raising the trustworthiness and internal validity by combination of one method of data collection. During the FGDs the participants were encouraged to speak freely and give their perspective about HIV risks factors and needs in HIV testing and prevention unique to their age group. Confidentiality was maintained and participants were strictly identified by serial numbers and not their names during the FGDs. The discussion was tape recorded for reference during analysis and report writing. Each FGDs session took about 60 minutes.

### 2.6. Data Management and Analysis

All written and digital information are kept in the strictest confidence and are inaccessible to anyone but members of the research team. Identifying information were removed from transcribed data and audio-recordings. Transcripts were entered into MAXQDA software program for coding and qualitative data analysis in a password protected computer to avoid unauthorized access. Following the transcription of the interviews, the researcher read and reread the transcripts several times and making personal notes and reflections.

Then the transcribed notes along with field notes were subjected to line-by-line analysis by author paying close attention to experiential claims and understandings of the participants regarding HIV risk and needs in testing and prevention information by the emerging adults in rural setting in Kenya. The key words, phrases and descriptions from the participants were collated in the process of identifying the codes in an iterative process. The convergence and divergence of data were noted in the process of developing the preliminary emergent themes. These themes were further interrogated and refined with a second reviewer from the research team.

### 2.7. Ethical Consideration

This study was reviewed and approved by the University of Wisconsin-Milwaukee’s Institutional Review Board (IRB) and Moi University/Moi Teaching and Referral Hospital Institutional Research and Ethics Committee (IREC). Written informed consent was obtained from all participants before the study began. Participants were informed of their rights, including the right to withdraw from the study at any time. The researcher reviewed the study’s informational sheet with each participant prior to beginning the interview to ensure understanding and informed consent. For participants under 18 years old, parental or guardian consent was obtained. Confidentiality was ensured by de-identifying all collected data and securely storing it. Participants received a $5 airtime voucher as a token of appreciation for their participation.

## 3.0. Results

### 3.1. Socio-Demographic Characteristics of the Participants

The socio-demographic characteristics for the participants is summarized in table 1 below.

**Table 1:** The Demographic Data for the Participants Interviewed (n=30).

| Variable | Categories of Variables | Count (X) | Percentage |
| --- | --- | --- | --- |
| Age (18-24) | 18 Years | 5 | 16.7 |
|  | 19 Years | 3 | 10.0 |
|  | 20 Years | 3 | 10.0 |
|  | 21 Years | 6 | 20.0 |
|  | 22 Years | 1 | 3.3 |
|  | 23 Years | 3 | 10.0 |
|  | 24 Years | 9 | 30.0 |
| Marital Status | Single | 25 | 83.3% |
|  | Married | 4 | 13.3% |
|  | Separated/Divorced | 1 | 3.3% |
| Number of Children | 0 | 23 | 76.7% |
|  | 1 | 6 | 20.0% |
|  | 2 | 1 | 3.3% |
| Employment Status | Self-Employed | 4 | 13.3% |
|  | Not employed | 12 | 40.0% |
|  | Student | 14 | 46.7% |
| Main Source of Income | Regular employment | 0 | 0.0% |
|  | Casual jobs | 12 | 40.0% |
|  | Support from family/friends | 14 | 46.7% |
|  | Self Employed | 4 | 13.3% |
| Average Income/Month | < Ksh 5000 | 22 | 73.3% |
|  | Ksh 5,001-10,000 | 7 | 23.3% |
|  | Ksh 10,001-15,000 | 1 | 3.3% |
| Financial Situation | Barely adequate | 13 | 43.3% |
|  | Inadequate | 17 | 56.7% |
| Level of Education | Primary | 1 | 3.3% |
|  | Secondary | 21 | 70.0% |
|  | Post Primary certificate Training | 5 | 16.7% |
|  | Associate Degree/Diploma | 1 | 3.3% |
|  | Bachelors | 2 | 6.7% |
| Currently in College | Yes | 15 | 50.0% |
|  | No | 15 | 50.0% |
| Current Living Situation | Rental a house/apartment | 9 | 30.0% |
|  | Family House | 17 | 56.7% |
|  | Spouse/Sexual partner | 2 | 6.7% |
|  | Student Hostel | 2 | 6.7% |
| HIV Status | Negative | 17 | 56.7% |
|  | Positive | 0 | 0.0% |
|  | Not ready to Disclose | 2 | 6.7% |
|  | Never tested | 11 | 36.7% |

We used the Modified Social Ecological Model (MSEM) framework to provide a structured analysis of the unique risk factors driving the HIV and AIDS epidemic among emerging male adults in rural Kenya. The MSEM is a health promotion model that addresses both individual behaviors and social-environmental factors, recognizing their combined impact on health outcomes. It outlines the complex interaction of multiple influences—Intra-personal/Individual, Interpersonal, Institutional, and Public Policy—each of which plays a role in HIV acquisition and prevention.

Our findings align with these MSEM levels, highlighting multiple barriers and facilitators across each category. The emerging male adult’s access to HIV prevention opportunities was examined under the major theme: “*Addressing the Unique Needs of Emerging Male Adults in HIV Prevention in Rural Settings”*. Within this major theme, initial codes were collated and grouped into four sub-themes, each corresponding to one of the MSEM framework levels. The major theme and subthemes reinforce that young men in rural settings face multiple interrelated challenges in accessing and utilizing HIV information and prevention services. Each sub-theme directly corresponds to a level within the MSEM framework, illustrating how individual behaviors, social networks, healthcare institutions, and policy environments collectively shape HIV prevention outcomes. This structured categorization ensures that prevention efforts address barriers holistically rather than in isolation, allowing for tailored interventions that respond to individual, social, institutional, and policy-level factors.

The young men participating in the focus group discussions (FGDs) and individual interviews reported several factors that heightened their risk for HIV infection. Table 2 below outlines the theme and subthemes mapped to the MSEM framework.

**Table 2.**
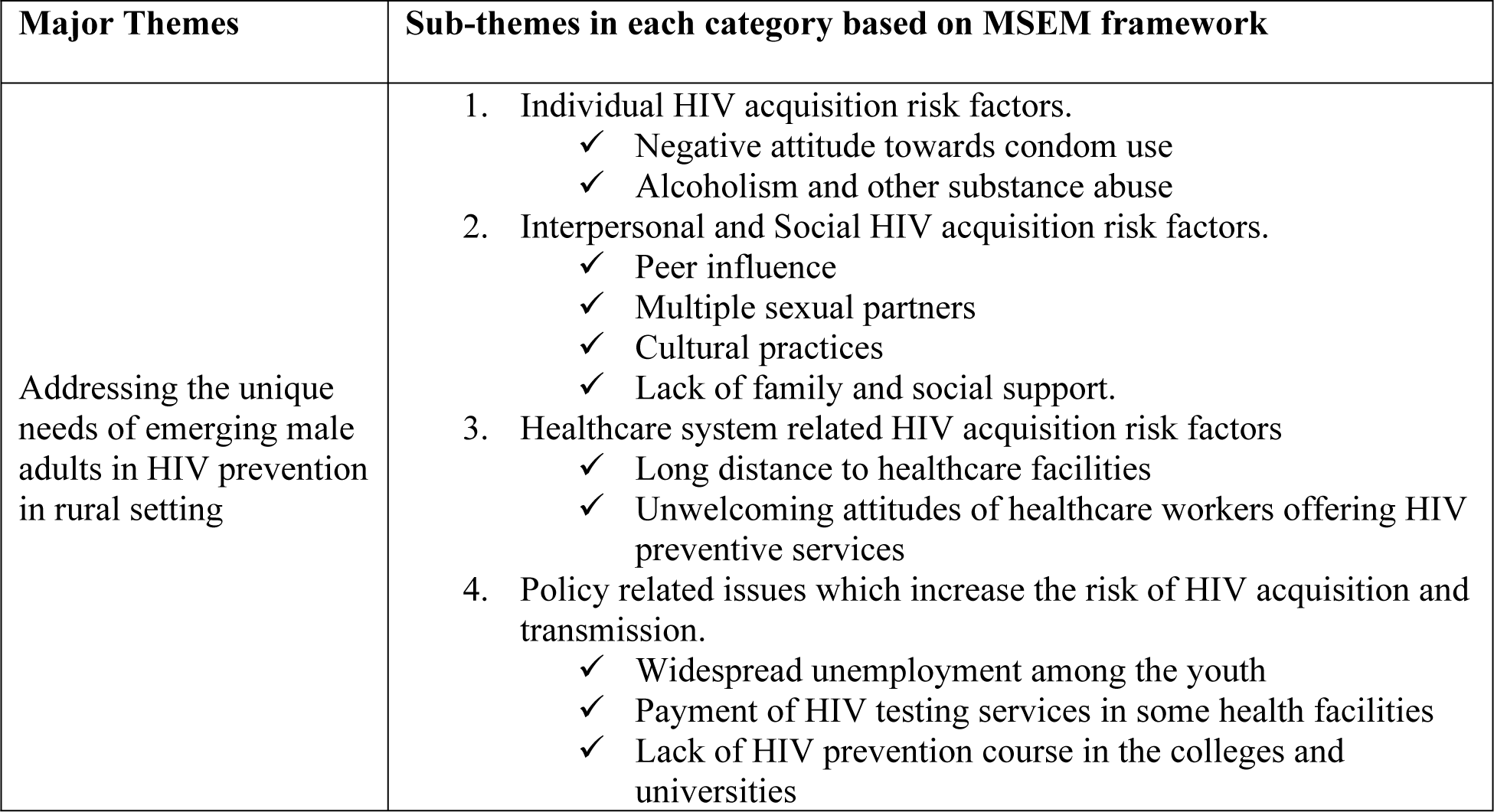
The Major Theme and Subthemes based on the MSEM Framework.

| Major Themes | Sub-themes in each category based on MSEM framework |
| --- | --- |
| Addressing the unique needs of emerging male adults in HIV prevention in rural setting | 1. Individual HIV acquisition risk factors.<br>✓ Negative attitude towards condom use<br>✓ Alcoholism and other substance abuse<br>2. Interpersonal and Social HIV acquisition risk factors.<br>✓ Peer influence<br>✓ Multiple sexual partners<br>✓ Cultural practices<br>✓ Lack of family and social support.<br>3. Healthcare system related HIV acquisition risk factors<br>✓ Long distance to healthcare facilities<br>✓ Unwelcoming attitudes of healthcare workers offering HIV preventive services<br>4. Policy related issues which increase the risk of HIV acquisition and transmission.<br>✓ Widespread unemployment among the youth<br>✓ Payment of HIV testing services in some health facilities<br>✓ Lack of HIV prevention course in the colleges and universities |

*3.1. Intra-personal/Individual HIV acquisition risk factors.*

### 3.1. Intra-personal/Individual HIV acquisition risk factors

The participants interviewed highlighted negative attitude towards condom use, alcoholism and other substance abuse as the major individual factors increased the risk for HIV transmission among the emerging adults in the rural areas. Some of the negative attitudes and perceptions reported by the participants towards condom use includes, condoms reduce pleasure, condom use as a sign of unfaithfulness. For many men interviewed, safety from HIV does not play a central role in their decision-making about condoms as one says. One participant during the individual interview stated:

*When I use a condom, my girlfriend will feel I am cheating or her so to avoid suspicious I rather not use it*.

Another participant during the FGDs said:

*My girlfriend recently told me that when we use the rubber (condom), it can easily slip and get lost in her uterus and we all got scared when we think of using it during the act (sexual intercourse)*.

Alcoholism and other substance abuse are another individual factor which predisposes young men to HIV infection. They gave many reasons as to why young people are engaging in drug abuse, the main one being frustrations arising from very unemployment rate currently in Kenya. Those who reported drinking alcohol expressed that they might forget to use a condom during sexual intercourse due to influence of alcohol. One participant in the interview stated:

*The reason why I resorted to drinking because I can’t secure any job even with by degree in procurement and supply chain management, and I am also sexually active, and I don’t even remember to use a condom when I am drunk*.

### 3.2. Interpersonal and social HIV acquisition risk factors

The participants mentioned the following interpersonal factors which increase the risk to HIV infection among the emerging adults in rural areas; peer influence and having multiple sexual partners, lack of family and social support including not going to churches. Faith-based teachings played a crucial role in shaping attitudes toward sexual health. Given the trust and community engagement of religious institutions, integrating faith-based organizations into HIV prevention programs could enhance intervention effectiveness, particularly in rural settings.

Collaborative efforts between public health initiatives and faith communities could help bridge gaps in HIV education and testing. Given the trust and community engagement of religious institutions, integrating faith-based organizations into HIV prevention programs could enhance intervention effectiveness, particularly in rural settings. One participant in the interview said:

> *I am a Christian from African Inland Church, and in our church, we usually have youth camps where a speaker in invited to speak about sexuality and sometimes even about HIV. These sessions emphasize about living a pure life and seek guidance from the lord to help overcome extramarital sexual temptations until after marriage is church. Personally, these teachings have been helpful*

Another participant during the FGDs explained the following about cultural practices common in their village.

> *Every December during long holidays teenage boys are initiated to men through circumcision and secluded teachings in the nearby forest. Most of these young men when they came out from seclusion, they are under peer pressure to engage in sexual intercourse with their girlfriends to proof that they are no longer children. This is how some get infected with HIV.*

Our in-depth interview showed that the sexual norms that prevailed in the families, communities and peer groups of the young men shaped their sexual behaviors and attitudes. One participant in the FGDs said that:

> *In this village, most of the men have multiple sexual partners…so having many partners is a normal thing. some men were in polygamous relationships with the consent of their partners.*

### 3.3. Healthcare system related HIV acquisition risk factors

Participants frequently mentioned lack of access to health care due to distance and unwelcoming comments from healthcare workers as a factor that contributed to differences in rural community HIV infection risk in that it prevented them from visiting the health centers and obtaining necessary health information that could be used to protect themselves from acquiring HIV and other STIs. For example, one participant during the FGDs stated:

> *It’s harder for us young people. I feel like our age groups in the urban areas have more access to doctors and information that they need because they live closer to the health facilities. In this village you must have to walk over 10 kilometers to the nearest health center.*

The visiting hours to health facilities were mentioned to be inconveniencing. Most of them reported that they engage in casual labor like construction and farming which takes the whole day and usually the visiting hours in all these health facilities and over when are they are free to visit for HIV preventive information. For example, one participant in the individual interviews lamented:

> *I have a major problem with our nearby dispensary vising hours, the 8AM-4PM will not work for hustlers like me because these are the hours, we are busy working, the hospitals to be considerate to us and set aside time outside normal working hours so that we can visit the facility for HIV testing and prevention information.*

### 3.4. Policy and laws that increase the risk for HIV acquisition and transmission

The participants reported some of the laws and policies which increases the risk to HIV acquisition include widespread unemployment among the youth, payment of HIV testing services and lack of HIV prevention course in the colleges and universities. All the young men interviewed were not having any formal employment and they raised their concerns and wanted the government to address the issues before it gets out of hand. The young men interviewed reported that unemployment has led to frustrations and alcoholism which in most cases lead to irresponsible sexual behavior. One participant during the interview said.

> *I graduated with Surveying course from a local Technical College in 2019. I am constantly looking for any form of employment, but I can’t find employment, it is getting harder everyday even to secure casual work. The government need to help create jobs for the youth. I am not choosy, I am ready for any kind of a job so that I can put food on the table.*

Another participant during FGDs session reported about payment for HIV self-testing kits as a discouraging issue for them to get HIV testing.

> *The HIV self-testing is popular among the young men. I like testing myself using the HIV self-test kit, but they sell it at Ksh 500, when you go to chemist, they sell it, yet the government says it is provided free of charge*

One participant in the interview expressed the view that the college education needs to be reviewed to include a general course about HIV and preventive measures. He reported that he just graduated from a local university recently, but he did not cover ant HIV related content in his entire university education.

> *I have a degree in education and in the entire university curriculum we were never taught HIV and AIDS content. The policy makers in higher education need to review the university education curriculum to include content in HIV and prevention.*

## 4.0. Discussion

The young men in the rural setting are faced with myriad of risk factors and challenges in accessing and utilizing HIV information and prevention services. The study was conducted in 2021. However, the barriers identified such as stigma, digital literacy, and lack of healthcare access remain highly relevant today. Limited digital literacy continues to be a barrier, preventing young men in rural areas from effectively utilizing mobile health interventions. Strategies such as community-based digital literacy training and simplified mobile health applications tailored to low-tech users could enhance engagement and accessibility. Digital health solutions require user-friendly designs and educational campaigns to enhance accessibility and engagement.

Recent studies on HIV prevention in rural Kenya confirm that these structural barriers persist, reinforcing the applicability of our findings. The young men reported individual, interpersonal, health system and policy level factors which heighten their risk for acquisition and speed of HIV infection in the rural communities in Kenya. The negative attitude towards condom use was common among the participants interviewed and posed as a major individual level risk factor for HIV acquisition and transmission. Several studies posits that correct and consistent use of a condom is needed for efficacious prevention in high prevalence settings particularly in SSA (12,13). This is consistent with findings from studies conducted in SSA which reported that even though AYA’s have good knowledge about condom use, they seldom use it correctly and consistently and still engaged in risky sexual behavior (13–15).

The HIV/AIDS misconceptions among the rural emerging males may be barriers to HIV prevention, thus it is important to create awareness and overcome misconceptions about HIV and AIDS in rural communities and changing young men’s HIV/AIDS misconceptions may promote men’s positive attitudes and beliefs in condom use and protect themselves from HIV (15–17). It is also important to develop a culturally appropriate HIV-prevention messages that address such beliefs and perception among the young people in order to have sustainable HIV prevention interventions.

Alcoholism and other substance abuse elevates their risk to acquisition of HIV because young men are unlikely to use a condom when they were under influence of alcohol or other substance of abuse. These findings are consistent with other studies conducted in SSA rural setting that showed that alcohol was perceived as a social lubricant by most AYA’s, which often led to risky sexual behaviors (18,19). The association between alcohol and substance abuse and HIV acquisition and transmission has been well documented as directly affect cognitive ability and judgement, which can lead to high-risk sexual behaviors, including unprotected, multiple sexual partners, and coercive sex (20,21).

The participants reported some cultural practices like rite of passage to adulthood through circumcision and secluded teachings increases the pressure to engage in sexual intercourse with their girlfriends to proof that they have transitioned to adulthood. It is well documented that sexual behaviors and cultural norms are interconnected, it is through culture that people learn how to behave and understand the world around them (22). In many cultural contexts, young men are taught from a very young age how to behave based on dominant notions of what it means to be a man in that context. As such, in some cultural context sexual risk-taking such as having multiple sexual partners and unprotected sex are perceived as normal behavior for men (18,22).

Some of the youth acknowledged that going to church and frequent teachings in the church that engaging in pre-marital sex is sin helped them stay away from risky sexual practices.

The church is well positioned to make important contributions to HIV prevention in SSA because they are more trusted by the indigenous population and are well placed to disseminate HIV and AIDS education messages (23–25). The church also provides some social support to people living with HIV and AIDS or even those engaged in alcoholism and drug abuse.

Our finding of a negative attitude among healthcare workers towards PLHIV is consistent with other studies and is associated with non-visits to HIV care and treatment centers.

Considering the sparsity of health services in rural areas of most of sub-Saharan Africa there is need to train health personnel in the importance of empathy towards patients, as well as engaging patients as partners in the HIV care process (26). Such negative attitudes will be barriers to service utilization by adolescents and young people and hampers the efforts to prevent HIV transmission (27,28). (We therefore call for a targeted effort toward alleviating negative attitudes toward youth and young people-friendly HIV and reproductive health service.

HIV self-test is popular among the young men interviewed. They reported that it is convenient and easy to use however they were disappointed that it is being sold in most outlets especially local pharmacies. The HIV self-test kits high acceptability among men has been reported in other studies in SSA. The majority of men showed a willingness to use HIV self-testing (HIVST) in studies conducted in Malawi, Tanzania, South Africa, and Kenya (29,30). The HIVST is highly popular among young men possibly because they engage themselves with HIV testing services without visiting health facilities. In addition to that, the HIVST model further empowers young people and sexually active individuals to be independent and have the option to choose the location and timing of the test and to control the disclosure of their results (31).

### Limitations

There are some limitations to this study. Methodologically, this is a qualitative study using a mixed method study might have yielded more interesting experiences in this age group of participants. This study took 6 weeks and data collection for a longer period and a longitudinal approach might yield more nuanced experiences from the participants.

Another limitation is that this study was conducted in rural Uasin Gishu in Western Kenya and will not include emerging male adults in urban areas therefore their views might not be captured. Nevertheless, while this study will not investigate the unique needs of emerging male adults in HIV prevention in urban setting, the findings from this study could still be transferable to urban populace. It is also justifiable to target the rural population because literature review reveals that there is limited research on HIV prevention and testing needs of adolescents and emerging male adults in rural settings in SSA and the few available target the young people living in urban towns and cities.

### Conclusion

In conclusion our findings showed the young men in the rural setting are faced with myriad of risk factors and challenges in accessing and utilizing HIV information and prevention services. Understanding these factors will inform the development of tailored prevention interventions and programs. This study underscores the need for innovative and contextually tailored interventions to address HIV prevention among emerging adults, ensuring accessibility while mitigating barriers such as digital literacy and stigma, including educational, social, policy, and health care systems that influence prevention knowledge, service use, and treatment options for youths.

## Data Availability

All data generated or analyzed during this study are included in this published article and its supplementary information files. The raw data supporting the conclusions of this manuscript will be made available by the authors, without undue reservation, to any qualified researcher. The qualitative data derived from in-depth interviews and focus group discussions are stored securely with the research team at University of Wisconsin-College Of Nursing. Due to the sensitive nature of the information collected and the privacy promises made to the participants during the consent process, the data are available from the corresponding author upon reasonable request and with approval from the University Of Wisconsin Institution Ethics Review Board. Researchers interested in accessing the data can contact the corresponding author, Dr. Augustine Kiplagat Bingat, via email at. Requests for access will be reviewed by the University of Wiscinsin Ethics Review Board] to ensure they comply with the ethical standards and privacy protections outlined in the consent agreements.

## Acknowledgments

We gratefully acknowledge the contributions of various individuals who supported this research but do not qualify for authorship. We would like to thank the community leaders in Ainabkoi Sub-County for their invaluable assistance in coordinating the participant recruitment process. Their insights and support were critical in facilitating our engagement with the study participants.

Special thanks to the field staff and volunteers who dedicated countless hours to ensuring the smooth running of data collection, and to the healthcare professionals in Uasin Gishu County who provided expert advice on our methodology and intervention strategies.

We also extend our appreciation to the participants themselves, whose willingness to share their personal experiences and perspectives has been essential to this study. Their contributions have not only enriched our research but also aim to enhance HIV prevention efforts within their communities.

Each individual named in this acknowledgment has given their consent to be mentioned.

## Notes

### Competing Interest Statement

The authors have declared no competing interest.

### Funding Statement

His research did not receive any specific grant from funding agencies in the public, commercial, or not-for-profit sectors. The authors conducted this study as part of their academic and professional responsibilities at their respective institutions, which provided the necessary support for the research, analysis, and preparation of the manuscript.

### Author Declarations

The research described has been reviewed and approved by the Ethics Review Board of University of Wisconsin Milwaukee. This study was conducted in accordance with the ethical standards of the responsible committee on human experimentation and with the Helsinki Declaration.

